# Treatment nonresponse in children receiving silver diamine fluoride for caries: A pilot study

**DOI:** 10.1101/2020.10.02.20204743

**Authors:** Ryan Richard Ruff, Bidisha Paul, Maria A Sierra, Fangxi Xu, Yasmi Crystal, Xin Li, Deepak Saxena

**Affiliations:** New York University College of Dentistry; New York University School of Global Public Health

## Abstract

*Objectives*: Silver diamine fluoride (SDF) is a nonsurgical therapy for the arrest and prevention of dental caries with demonstrated clinical efficacy. Approximately 20% of children receiving SDF fail to respond to treatment. The objective of this study was to develop a predictive model of treatment nonresponse using machine learning. *Methods*: An observational pilot study (N=20) consisting of children with and without active decay and who did and did not respond to silver diamine fluoride provided salivary samples and plaque from infected and contralateral sites. 16S rRNA genes from samples were amplified and sequenced on an Illumina Miseq and analyzed using QIIME. The association between operational taxonomic units and treatment nonresponse was assessed using lasso regression and artificial neural networks. *Results*: Bivariate group comparisons of bacterial abundance indicate a number of genera were significantly different between nonresponders and those who responded to SDF therapy. No differences were found between nonresponders and caries-active subjects. Prevotella pallens and Veillonella denticariosi were retained in full lasso models and combined with clinical variables in a six-input multilayer perceptron. *Discussion*: The acidogenic and acid-tolerant nature of retained bacterial species may overcome the antimicrobial effects of SDF. Further research to validate the model in larger external samples is needed.

## 1 Introduction

Dental caries, a polymicrobial disease caused by a diverse bacterial ecosystem where acid produced by acidogenic and aciduric bacteria in direct contact with tooth surfaces erodes the enamel and dentin of the tooth [1], is the most prevalent childhood disease in the world [2, 3]. Untreated dental caries affects more than 20% of children in the United States and over 50% of children have ever experienced caries, higher amongst low-income and minority children [4, 5, 6], populations typically characterized by lower dental service utilization [7]. To address this unmet need, school-based caries prevention programs can supplement traditional care, increasing access to dental services and reducing oral health inequities [8, 9].

Silver diamine fluoride (SDF) is a liquid therapeutic agent that combines the antibacterial effects of silver and the re-mineralizing effects of fluoride [10] with proven clinical efficacy in arresting approximately 80% of caries lesions in enamel and dentin [11]. The use of SDF for arresting cavitated caries lesions in primary teeth is recommended by the American Association of Pediatric Dentists and the American Dental Association [12] with growing utilization in pediatric dental programs across the United States.

The mechanisms associated with refractory growth of caries initially treated using SDF remain unclear. In pragmatic studies, this phenomenon of nonresponse to the therapeutic effect of SDF has been observed in children with and without prior caries experience, in those living in both fluoridated and non-fluoridated communities, and was not related to sex or age [13]. The presence of plaque in lesions resistant to the caries arresting action of SDF suggests that the microbiome composition of the individual oral cavity and/or the specific caries site may be a strong determinant in the preventive action of SDF [11, 14]. Silver ions have demonstrated an antimicrobial effect against specific bacteria and the development of new biofilms [15, 16]. However, most studies with cariogenic bacteria have been conducted in in-vitro biofilm models with one or more species while the human biofilm is composed of multitude of species, the interaction of which may influence the effect of SDF in preventing caries.

We conducted a pilot study to explore the innate commonalities between the microbiomes of children that may increase susceptibility or resistance to caries and impact the effectiveness of SDF therapy. Study objectives included determining whether (1) caries prone or resistant children harbor significantly different oral microbiota; (2) microbiomes are more or less resistant to preventive agents; and (3) these data can be used to predict treatment nonresponse. Results for objectives (1) and (2) were previously reported. In this paper, we present preliminary results supporting objective (3).

## 2 Methods

This study received approval from the New York University School of Medicine Institutional Review Board (#s19-00692, “The role of the oral microbiome in predicting disparities in caries and responsiveness to caries prevention: An observational pilot study”, 23 June 2019) and is reported according to STROBE checklist.

### 2.1 Design and participants

This was an observational pilot study conducted from June to November 2019. Twenty children aged 5-13 years were recruited from the New York University College of Dentistry dental clinic at the time of their routine visit. Healthy subjects were enrolled if they had untreated dental caries (N=5), were caries free (N=5), were scheduled to receive silver diamine fluoride during their current visit (N=5), or had previously received SDF and presented with re-occurrence of caries (N=5). Children in each group were unique and did not crossover. Excluded subjects were those who had antibiotic therapy in the previous two months or those who could not follow the instructions to collect the saliva samples due to mis-behavior. All study participants were of Hispanic/Latino origin. Each subject provided parental informed consent and child assent.

### 2.2 Sample collection

Untreated dental caries or re-occurrence of caries was determined by subjects presenting with visible cavitated lesions corresponding to an International Caries Detection and Assessment System (ICDAS) score of 3 or higher. Patients received silver diamine fluoride if they presented with visual cavitated lesions consisting of an ICDAS score of 4 or higher. Biological samples were collected once from each of the twenty study participants in a single visit. All subjects received a visual-tactile oral examination and provided unstimulated saliva. For children with caries, plaque samples were taken from caries sites and from the buccal surface of the second primary molar on the opposite side of the treated or untreated cavity. Children without caries provided supragingival plaque samples from the buccal surface of the second maxillary molar on both sides of the mouth. Supragingival plaque was collected with a single stroke using a sterile Gracey mini curette. Samples were placed in separate pre-barcoded micro centrifuge tubes with transport buffer and placed in a portable freezer. All biological samples were collected by a licensed dentist.

Sociodemographic and other clinical data for participants were obtained using pre-study questionnaires or were extracted from electronic health records. Data included subject age, sex, ethnicity, the number of hours since the last meal was consumed and since the last time the subject brushed his or her teeth, visible plaque index score, and caries history. While there was no control over the time of the last meal or their last brushing, however this data were collected and both were at least one hour in all subjects.

### 2.3 Microbiome analysis

Bacterial genomic DNA was extracted using QIAamp PowerFecal kit (Qiagen). DNA purity was verified using NanoDrop 2000 spectrophotometer (Thermo scientific) and quantified fluorometrically using Quant-iT PicoGreen assay (Invitrogen). Each of the samples were further diluted or concentrated to 10ng/ul. The variable V4 region of the bacterial rRNA gene was amplified in duplicates using barcoded forward and reverse primers 341F (5-CCTACGGGNGGCWGCAG-3) and 805R (5-GACTACHVGGGTATCTAATCC-3), each with overhang adapter sequences (IDT). PCR was done using 2x Kapa HiFi Hotstart ReadyMix DNA polymerase (KapaBiosystems). Reactions were verified by agarose-gel electrophoresis and amplicons were purified using AMPure XP beads. A second PCR was performed using dual indices from Illumina Nextera XT index kits (Illumina) followed by second purification with AMPure XP beads. The amplicons were further quantified by PicoGreen assay and diluted to 4nm. The samples were then pooled together, denatured, and sequenced on Illumina MiSeq platform (paired end, 2*300 bp).

### 2.4 Data analysis

Descriptive statistics (means, standard deviations, and proportions) were calculated for clinical and sociodemographic data. Any differences across study groups were assessed using *χ*^2^ tests for categorical variables and analysis of variance for continuous variables. T-tests with multiple comparison adjustments were used to detect any simple bivariate differences across the relative abundance of OTUs. Adjusted models for the association between taxonomic units and treatment nonresponse was then assessed using lasso regression [17, 18]. Study groups were collapsed into those who were nonresponders to SDF and all other subjects (caries active, caries free, and responders) due to small sample sizes in the pilot. All available OTUs were used as potential predictors with a single outcome consisting of two classes (non-responder versus all other study participants). Analysis used k-fold cross-validation to produce values for *λ*, after which a logistic lasso was performed. Coefficients of retained variables were saved and plots were produced for coefficient change across *λ* values.

Retained OTUs and clinical indicators for time elapsed since prior meal and brushing, age, and VPI were used as input variables for an artificial neural network. The included OTUs were first normalized using the function (*x – min*(*x*))*/*(*max*(*x*) – *min*(*x*)). A multilayer perceptron consisting of five hidden nodes, six inputs, and one output was constructed and computed using backpropagation. Predictions were compared to observed values but due to the small sample size no test set was used for validation.

Analysis was conducted in R v3.6.3 (http://www.r-project.org) using the glmnet and neuralnet packages. Statistical significance was set at 0.05.

## 3 Results

The average age of study participants was 9.1 (SD=2.15) with approximately 55% of the sample consisting of females (Table 1). Sex was unequally distributed across treatment groups (e.g., 20% female in caries active and 80% in SDF groups). The average elapsed time since the previous meal and the last toothbrushing was 4.35 (SD=3.62) and 3.25 (SD=1.97) hours, respectively. The average visible plaque index (VPI) score across the study sample was 2 (SD=0.56).

**Table 1:** Clinical and sociodemographic data (N=20)

|  | **All** | **Caries active** | **Caries free** | **Non-responders** | **SDF** |
| --- | --- | --- | --- | --- | --- |
| **Variable** | Mean (SD) | Mean (SD) | Mean (SD) | Mean (SD) | Mean (SD) |
| Age | 9.1 (2.15) | 9.4 (2.7) | 9.2 (1.48) | 9.8 (2.59) | 8 (1.87) |
| Time last meal | 4.35 (3.62) | 7 (4.69) | 2.6 (1.67) | 3.2 (2.17) | 4.6 (4.28) |
| Time last brush | 3.25 (1.97) | 3 (1.58) | 2.8 (2.39) | 4 (2.35) | 3.2 (1.92) |
| VPI | 2 (0.56) | 2 (0.71) | 1.8 (0.84) | 2 (0) | 2.2 (0.45) |
| Sex (females) % | 0.55 | 0.2 | 0.6 | 0.6 | 0.8 |

Bivariate group comparisons for abundance of specific genera in non-responders (Table 2) indicates that Oribacterium, Prevotella, Aggregatibacter, Fusobacterium, Peptostreptococcaceae, and Campylobacter were significantly different comparing non-responders to SDF-treated subjects. When compared to caries-free subjects, only Absconditabacteria and Peptostreptococcus were significantly different. There were no significant differences between non-responders and caries-active individuals. In full lasso models, two species were retained (Table 3) at the lambda minimum: Pallens (genus Prevotella) and Denticariosi (genus Veillonella). The coefficient path showing changes in estimates for OTUs for included lambdas is shown in Figure 1.

**Table 2:** Specific genera comparisons with non-response

| Group comparison | Taxa | p-value |
| --- | --- | --- |
| SDF | Oribacterium | 0.02695256 |
|  | Prevotella | 0.02662165 |
|  | Aggregatibacter | 0.02860014 |
|  | Fusobacterium | 0.02969937 |
|  | Campylobacter | 0.01704905 |
|  | Peptostreptococcaceae_[XI][G-9] | 0.0385394 |
| Caries Free | Absconditabacteria_(SR1)_[G-1] | 0.02393104 |
|  | Peptostreptococcus | 0.03700165 |
| Caries Active | N/A |  |

**Table 3:** Retained and potentially relevant OTUs for treatment nonresponse

| Retained | Phylum | Family | Genus | Species |
| --- | --- | --- | --- | --- |
|  | Bacteroidetes | Prevotellaceae | Prevotella | pallens |
|  | Firmicutes | Veillonellaceae | Veillonella | denticariosi |
| **Non-retained** | Bacteroidetes | Prevotellaceae | Prevotella | multiformis |
|  | Fusobacteria | Leptotrichiaceae | Leptotrichia | NA |
|  | Actinobacteria | Micrococcaceae | Rothia | dentocariosa |

**Figure 1:**
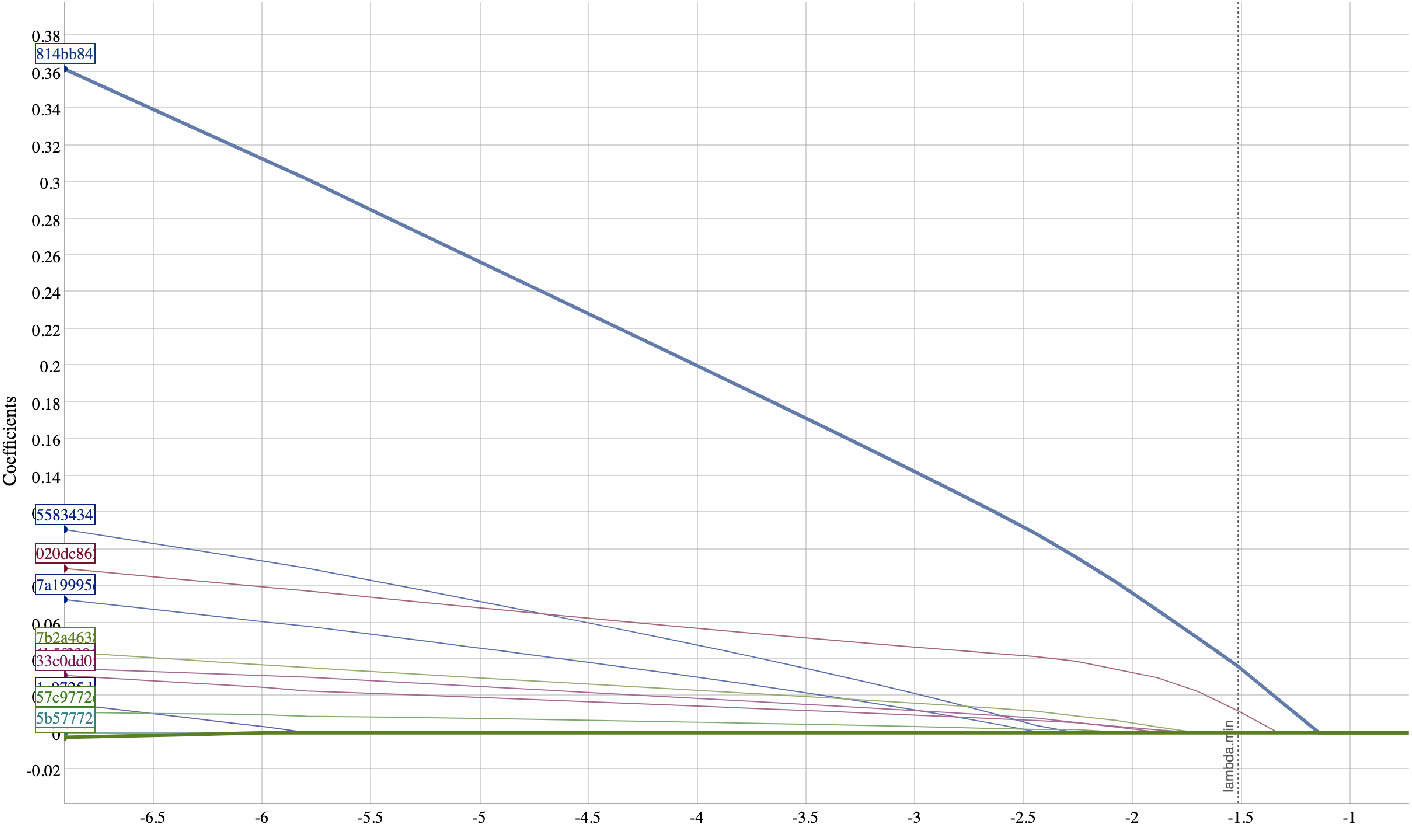
Coefficient path of OTUs at different values of lambda

At smaller estimates of lambda, additional OTUs were present (species Multiformis, Dentocariosa of the Rothia genus, and the genus Leptotrichia). Given the small sample size, these OTUs may be relevant in future analyses. An interactive graph of the coefficient path is available in a public GitHub repository (www.github.com/ryanruff/nonresponse). The network topology map for the two retained species and clinical indicators is shown in Figure 2. Associated weights for node connections (black lines) and bias (blue lines) are available in the GitHub repository.

**Figure 2:**
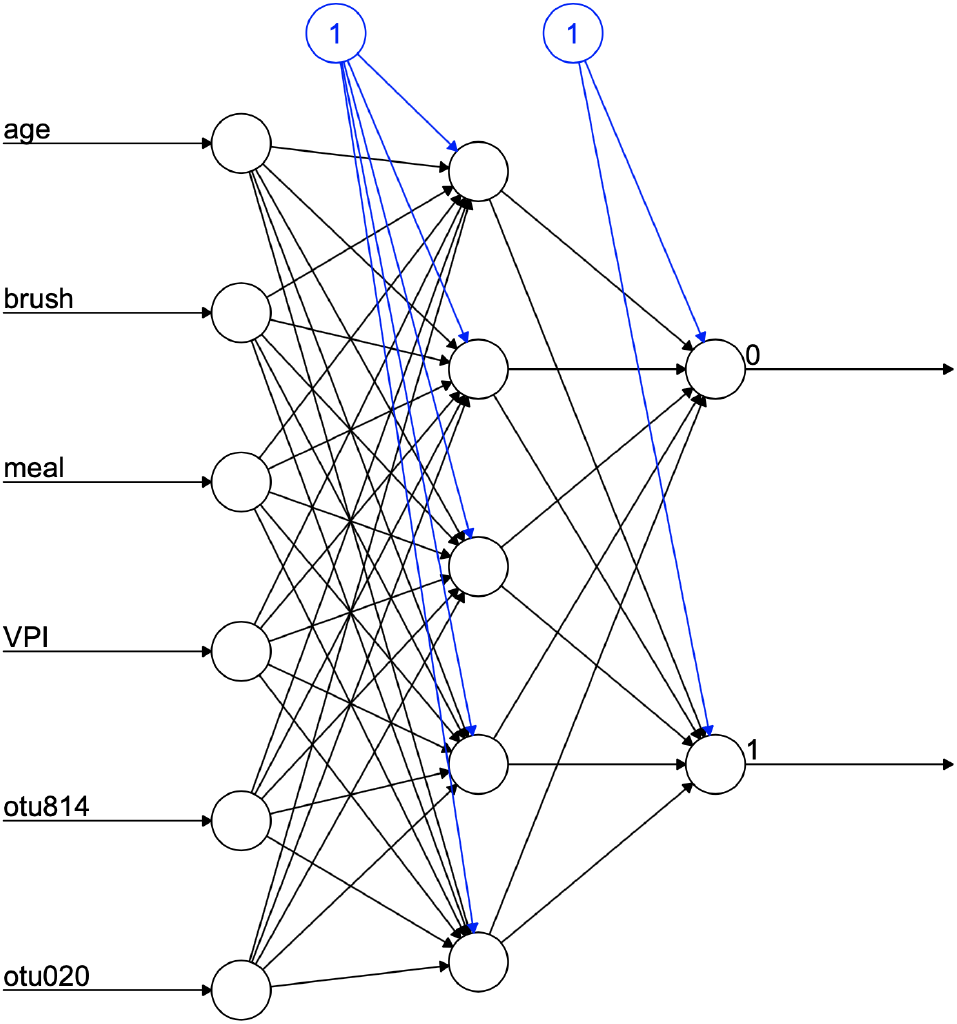
Network topology without weights

## 4 Discussion

Silver diamine fluoride combines the antibacterial behavior of silver with the remineralization properties of fluoride that creates an unfavorable environment for collagen degradation, and can inhibit the progression of dental caries [19]. We previously reported that there are distinct microbiota within plaque and saliva samples that can successfully differentiate between nonresponders to SDF and other groups, though no differences were found in microbial composition between infected or contralateral sites. In this study, our approach considered two methods to yield a predictive model for treatment nonresponse using a sample of patients who had previously received silver diamine fluoride for dental caries. While research using machine learning in oral epidemiology is limited, previous studies have successfully applied ML-approaches to predict malodor using salivary microbiota [20], hypertension from gingival inflammation [21], oral cancer [22], oral poliovirus vaccine immunogenicity [23], temporomandibular joint osteoarthritis, and radiographic detection of tooth and dental restorations [24]. Notably, a recent review concluded that artificial intelligence, particularly machine learning and deep learning, can use data from the oral microbiome to predict systemic disease [25].

Non-responders were compared to children with and without active caries, as well as those who have responded to the therapeutic effects of SDF. Interestingly, bivariate results indicate no differences in bacterial abundance between those that fail to respond to SDF and those with untreated carious lesions, and the greater number of differences were restricted to the SDF-treated group. Results using lasso regression indicated that two bacteria, Prevotella pallens and Veillonella dentocariosa, may be useful in predicting treatment nonresponse. Prevotella is proteolytic, saccharolytic and acid-tolerant [26], an overabundance of which has been associated with dental caries [27], in particular P. pallens [28]. The genus Veillonella consists of small strictly anaerobic gram-negative cocci that have been associated with severe early childhood caries [29]. Veillonella relies on lactate and other organic acid as its nutrient source and poorly adhere to host tissue. However, they are known to coaggregate with Streptococcus species such as S.mutans in biofilm formation in oral cavity [30]. These acidogenic and acid-tolerant bacterial species may overcome the antimicrobial effects of SDF, though the mechanistic action of this is still unknown.

Other species that were initially associated but were not retained at the lambda minimum included Prevotella multiformis, Rothia dentocariosa and the genus Leptotrichia. Prevotella multiformis, first isolated from oral gingival plaque, are gram-negative bacilli or cocci that produce acid from glucose, lactose, sucrose, glycerol, D-mannose, D-raffinose [31]. Rothia dentocariosa is a gram-positive bacteria with a variable morphology ranging from coccoid to branched filamentous, generally grows in aerobic and microaerophilic conditions producing acid from glucose, sucrose, maltose and glycerol [32], and has been previously associated with periodontal inflammatory disease. Notably, several gram-positive bacteria produce lipoproteins that induce TLR-2 mediated inflammation. Thus, Rothia denticariosa may be acting in concert with other gram-positive bacteria to invoke host-immune response and regulate inflammatory effects in dental caries [33]. Bacteria belonging to genus Leptotrichia are non-motile facultative anaerobe gram-negative bacilli that are found in oral cavity. Similar to streptococcus mutans, they ferment carbohydrates and produce organic acids such as lactic acid, and traces of acetic, formic and succinic acid. As S. mutans adheres to tooth surfaces and produces lactic acid causing demineralization of tooth enamel, Leptotrichia may us a similar mechanism to contribute to dental caries [34]. Further study on the potential of action of these bacteria in overcoming the effect of SDF is warranted.

The value of the coefficients from lasso models and neural networks are less important than the predictive power of the model and are thus unreported, though they can be extracted from supplementary figures. While the small sample size from this pilot study prohibited out of sample validation for testing purposes, our approach produced a restricted function for classifying whether a subject receiving silver diamine fluoride would appropriately respond to treatment. It is important to note that our results, as would be the case with any regularization approach, are only one of many possible models that could be useful in predicting nonresponse. Retained OTUs do not necessarily mean that those not included in the model are irrelevant. Regardless, this preliminary investigation has identified variables that are correlated with those predictive of treatment nonresponse, which may be useful in making predictions in larger, independent datasets.

The clinical definition of nonresponse to silver diamine fluoride is not yet fully explicated. In this study, any subject previously treated with SDF that presented with new incidence of decay was classified as a non-responder. However, an alternative classification might be to group individuals by the severity of nonresponse and the importance of individual operational taxonomic units might depend, in part, on the chosen clinical definition. Finally, independent variables for this study were limited solely to specific clinical indicators and OTUs produced by saliva and plaque samples taken from carious lesions and contralateral sites. Further research with more sophisticated data can expand upon the models and the neural network estimated in this study by incorporating data on dietary intake and hygiene behaviors, which are likely to confound the microbiome and nonresponse relationship.

### 4.1 Funding

This study was funded in part by awards from the New York University Grant Support Initiative (#RA633, Ruff PI) and the National Institute of Dental and Craniofacial Research (#R56DE028933, Ruff and Saxena PIs). The content is solely the responsibility of the authors and does not necessarily reflect the official views of the National Institutes of Health, New York University, the New York University College of Dentistry, or the New York University School of Medicine.

### 4.2 Contributions

RRR conceived and developed the study, performed statistical analysis, and wrote the manuscript. YC contributed to study development, enrolled subjects, collected samples, and wrote the manuscript. BP, MAS, and FX analyzed study samples and wrote the manuscript. DS contributed to study development and wrote the manuscript. All authors contributed equally.

## Data Availability

Code and models are available at a GitHub repository (www.github.com/ryanruff/nonresponse). Data available upon request and review.

